# Modelling, Simulations and Analysis of the First COVID-19 Epidemic in Shanghai

**DOI:** 10.1101/2021.06.20.21259203

**Authors:** Lequan Min

## Abstract

To date, over 178 million people on infected with COVID-19. It causes more 3.8 millions deaths. Based on a previous symptomatic-asymptomatic-recoverer-dead differential equation model (SARDDE) and the clinic data of the first COVID-19 epidemic in Shanghai, this paper determines the parameters of SARDDE. Numerical simulations of SARDDE describe well the outcomes of current symptomatic individuals, recovered symptomatic individuals, and died individuals, respectively. The numerical simulations suggest that both symptomatic and asymptomatic individuals cause lesser asymptomatic spread than symptomatic spread; blocking rate of about 95.5% cannot prevent the spread of the COVID19 epidemic in Shanghai. The strict prevention and control strategies implemented by Shanghai government is not only very effective but also completely necessary. The numerical simulations suggest also that using the data from the beginning to the day after about 19 days at the turning point can estimate well the following outcomes of the COVID-19 academic. It is expected that the research can provide better understanding, explaining, and dominating for epidemic spreads, prevention and control measures.

## 1 Introduction

In December 2019, a novel coronavirus-induced pneumonia (COVID-19) broke out in Wuhan, Hubei. Now over 178 million people on infected with COVID-19. It causes more 3.8 millions deaths. The COVID-19 epidemics affect more than 220 countries and regions including Antarctica. One of the reasons of such a tragedy is that people in some countries do not pay attention to theoretical analysis and estimations for COVID-19 epidemics.

In fact mathematical models for epidemic infectious diseases have played important roles in the formulation, evaluation, and prevention of control strategies. Modelling the dynamics of spread of disease can help people to understand the mechanism of epidemic diseases, formulate and evaluate prevention and control strategies, and predict tools for the spread or disappearance of an epidemic [1].

Since the outbreak of the COVID-19 epidemic in Wuhan, many scholars have published a large numbers of articles on the modeling and prediction of COVID-19 epidemics (for examples see [2–9]). However it is difficult to describe well the dynamics of COVID-19 epidemics. In a Lloyd-Smith et al’s paper, it described nine challenges in modelling the emergence of novel pathogens, emphasizing the interface between models and data [10].

In January 20, two Shanghai people were confirmed to be infected with COVID-19. Thus has caused the first wave COVID-19 epidemic in Shanghai. A total of 339 locally diagnosed cases were reported during the first wave COVID-19 epidemic. After 103 days, 332 COVID-19 patients were cured, and 7 patients died. The medical personnel has realized the zero infection.

Using one symptomatic-asymptomatic-recoverer-dead differential equation model (SARDDE) [11,12] and clinic data [13], this paper determines the parameters of the SARDDE and simulates the dynamic of the first COVID-19 epidemic in Shanghai [13]. The numerical simulations describe well the practical outcomes ([13]) of current infected symptomatic individuals, recovered infected symptomatic individuals, and died infected individuals.

The rest of this paper is organized as follows. Section 2 introduces the SARDDE and related theories. Section 3.1 implements the dynamic simulations of SARDDE to describe the data of the first COVID-19 epidemic in Shanghai. Section 3.2 states analysis and discussions; two virtual simulation examples are implemented to emphasize the importance of strict control measures and long terms’ estimation to epidemic spreading. Conclusions are given in Section 4.

## 2 SARDDE Model and Dynamic Properties

### 2.1 SARDDE Model [11, 12]

For SARDDE model, there are four states. *I*(*t*), *I*_*a*_(*t*), *I*_*r*_(*t*), *I*_*ra*_(*t*) and *D*(*t*) represent the fraction of current symptomatic infected individuals, and current asymptomatic but infected individuals, cumulative recovered symptomatic infected individuals, cumulative recovered asymptomatic but infected individuals and cumulative died individuals, respectively. The transition among these states is governed by the following rules (Flowchart of the rules is shown in Fig.1, where *S* represents susceptible population.).

**Figure 1:**
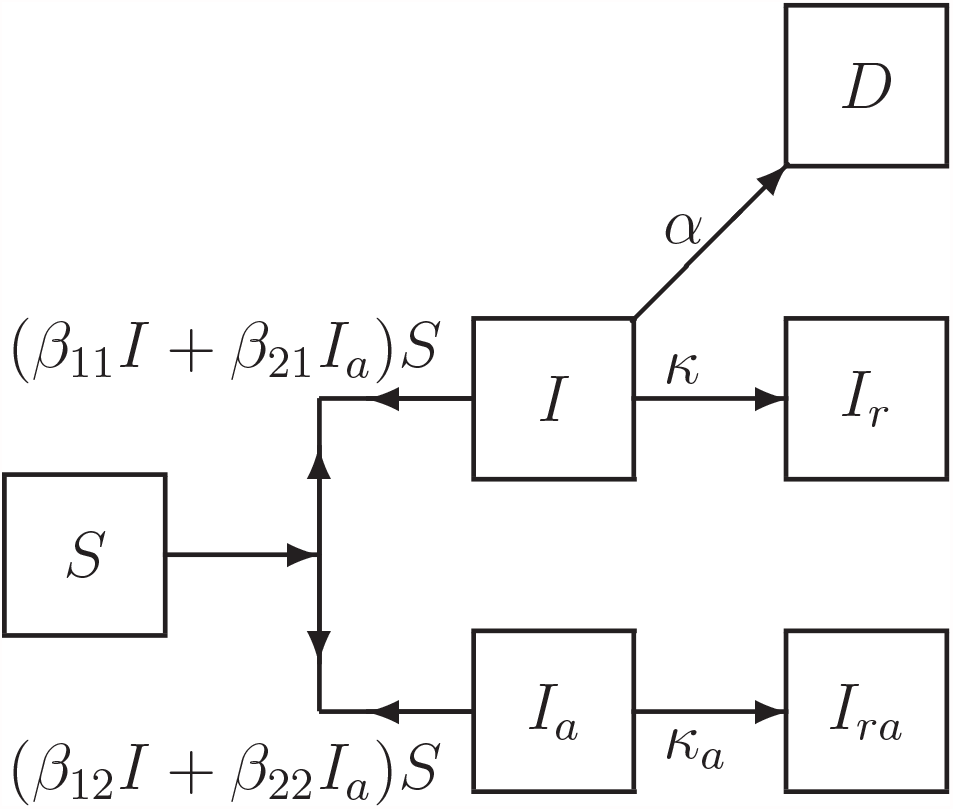
Flowchart of disease transmission among susceptible population *S*, current symptomatic infected individuals *I*, current asymptomatic but infected individuals *I*_*a*_ recovered symptomatic infected individuals *I*_*r*_, recovered asymptomatic but infected individuals *I*_*ra*_, and died individuals *D*.

First, the symptomatic infected individuals (*I*) and the asymptomatic but infected individuals (*I*_*a*_) infect the susceptible population (*S*) with transmission rates of *β*_11_ and *β*_21_, respectively, making *S* become symptomatic infected individuals, and with transmission rates of *β*_12_ and *β*_22_, respectively, making *S* become asymptomatic individuals. Then, a symptomatic individual is cured at a rate *κ*, an asymptomatic individual returns to normal at a rate *κ*_*a*_. An infected individual dies at a rate *α*. Here all parameters are positive numbers. Assume that the dynamics of an epidemic can be described by *m* time intervals. At *i*th interval, the model has the form:

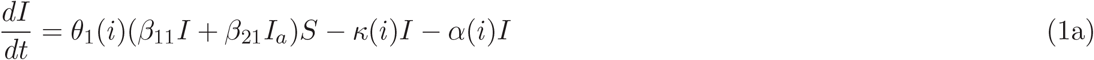

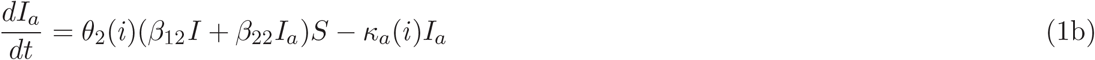

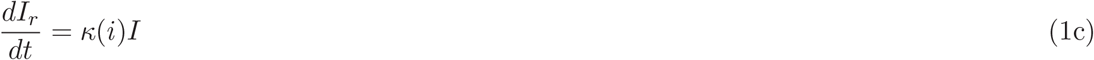

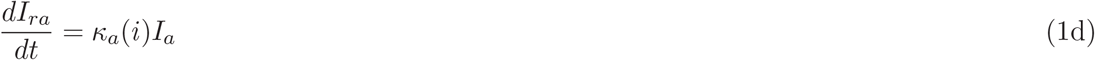

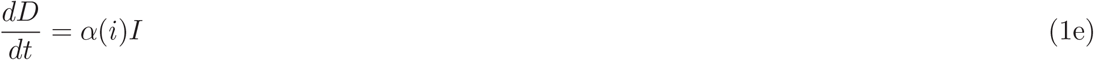

where (1 − *θ*_1_(*i*)′*s*) and (1 − *θ*_1_(*i*)′*s*) (*i* = 1, …, *m*) represent the blocking rates to symptomatic and asymptomatic infections, respectively.

The equation (1) has a disease-free equilibrium:

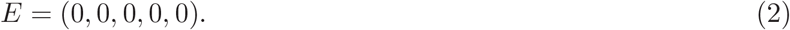

### 2.2 Two Theorems

The stability of system (1) is determined by the first two equations (1a) and (1b). Denote in (1a) and (1b)

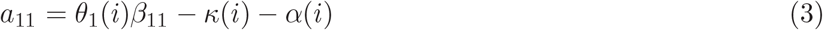

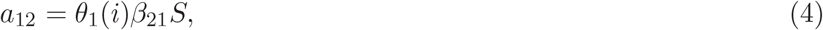

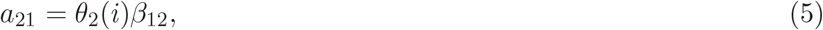

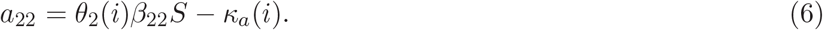

Then we have the following:

#### Theorem 1

*([11, 12]) Suppose that a*_11_, *a*_12_, *a*_21_ *and a*_22_ *are defined by (3)-(6) then the disease-free equilibrium E of system (1) is globally asymptotically stable if, and only if, the following inequalities hold:*

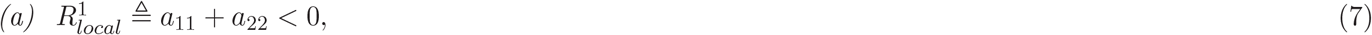

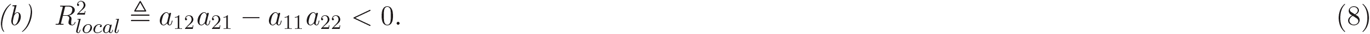

#### Theorem 2

*([11, 12]) If system (1) satisfy the following inequalities*

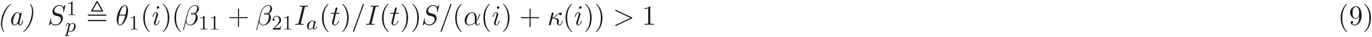

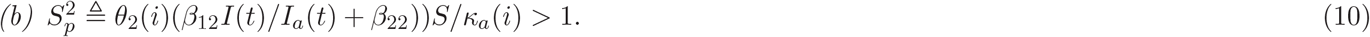

*then a disease transmission will occur*.

## 3 Applications

Based on the reported clinical COVID-19 epidemic data from January 20 to May 2, 2020 in Shanghai [13], this Section will simulate the dynamics of the first COVID-19 epidemic in Shanghai. Numerical simulations and drawings are performed by using MATLAB software programs.

The reported clinical data on current confirmed infection cases, and the reported clinical data on recovered cases of the COVID-19 epidemic in Shanghai [13] are shown in Figs. 2(a)-2(c). The number of current symptomatic infected individuals is showed in Fig3(a) by circles. The numbers of cumulative recovered symptomatic infected individuals, and cumulative died infected individuals are showed in Fig3(b) by circles and stars respectively.

**Figure 2:**
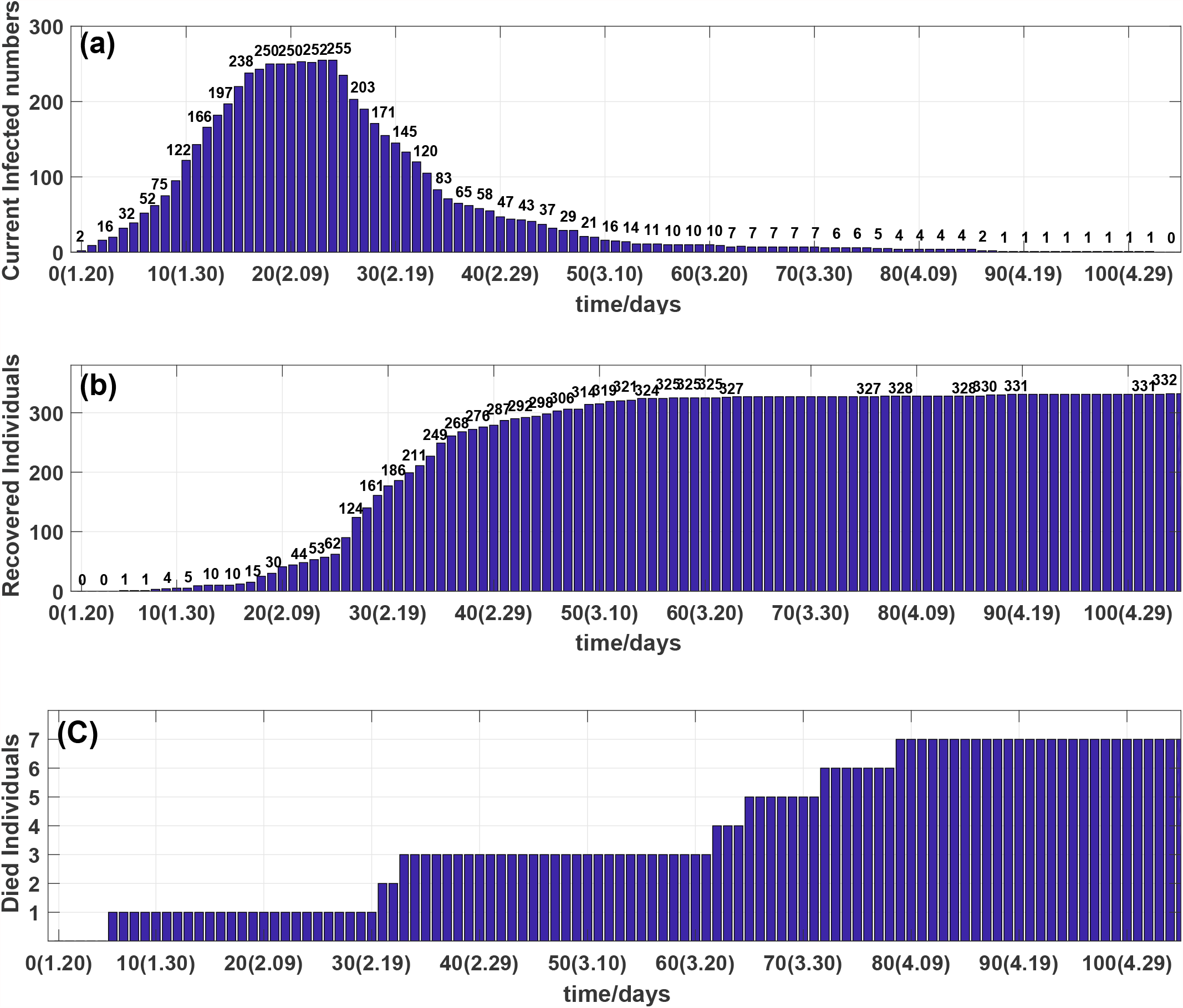
(a) Outcome of the number of current infected individuals. (b) Outcome of the number of cumulative recovered individuals. (c) Outcome of the number of cumulative died individuals.

**Figure 3:**
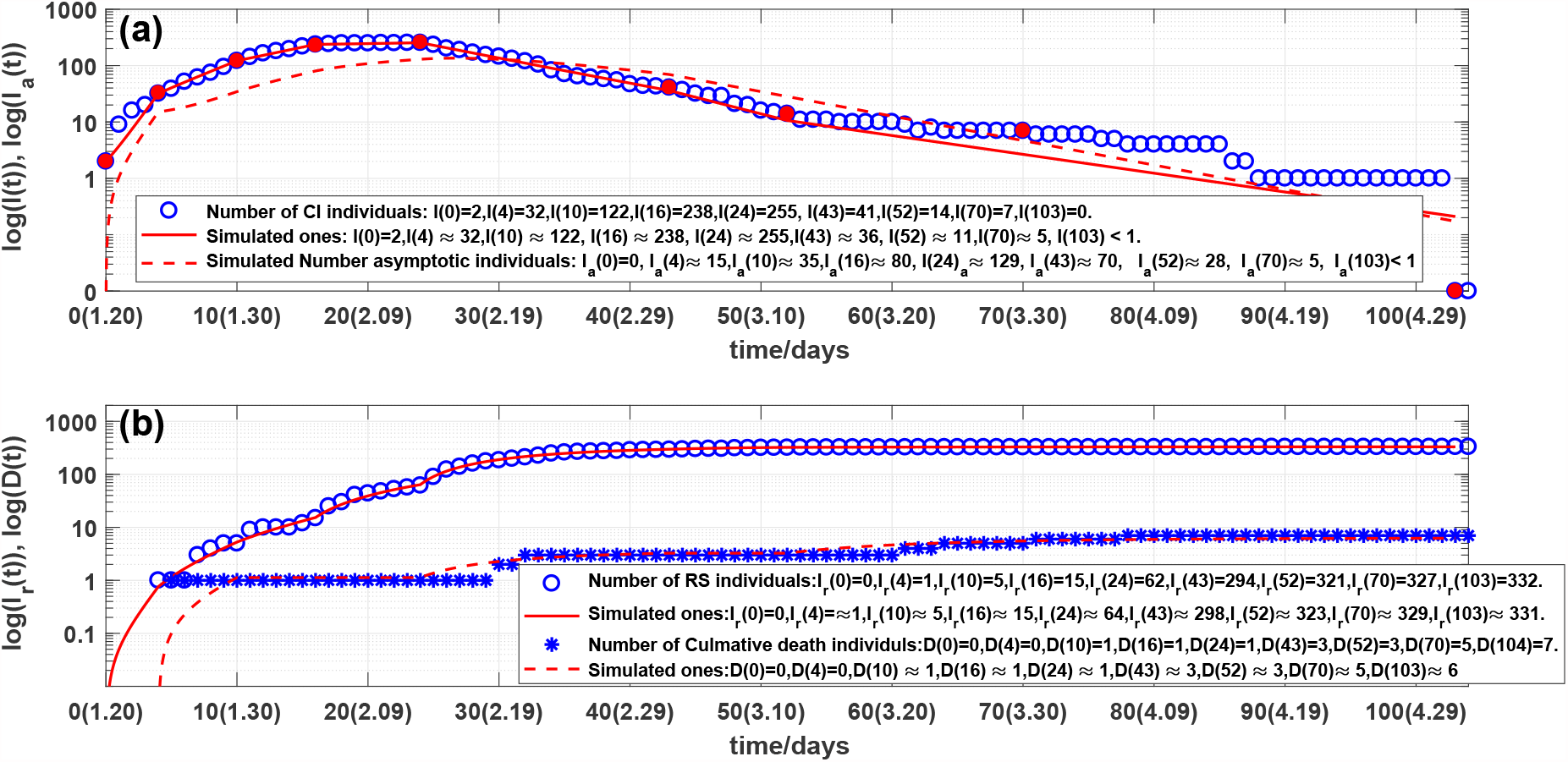
(a) Outcome of the number of current symptomatic individuals, representing by circles. Solid line and dash line are outcomes of stimulated current symptomatic individuals and stimulated current asymptomatic individuals of system (1). (b) Outcomes of the numbers of cumulative recovered symptomatic and died individuals, representing by circles and stars, respectively. Solid line and dash line are corresponding simulations of system (1).

The number of current infected individuals was risen rapidly in the first 4 days (see Fig. 2(a)). The number of current infected individuals reached the highest 255 on the day 24th, February and then declined rapidly (see Fig. 2(a) and Fig.3(a)).

Observe from the Figs. 3(a) and 3(b) that the overall changes in the numbers of current confirmed symptomatic and asymptomatic cumulative recovered individuals, and cumulative died individuals are not subject to the law of exponential changes, but the data can be approximated in good agreement with 8 straight lines in log scale (see Fig. 3(a)). This phenomenon can be explained as: different medical measures prevention and control strategies have been adopted at the different 8 time intervals. Therefore the *i* in SARDDE model (1) should be chosen as *i* = 1, 2, …, 8.

### 3.1 Simulation and prediction of the first COVID-19 epidemic in Shanghai

First it needs to determine the parameters *κ*(*i*), *κ*_*a*_(*i*) and *α*(*i*).There are different methods for calculating the recovery rate *κ*(*i*) in a specific time interval. Denote *s*_1_(*i*) and *s*_2_(*i*) to be the days that the old patients and the new patients stayed in the hospital during *i*th time interval. Denote *R*(*i*) and *d*(*i*) to be the numbers of the recovered patients and died patients during *i*th time interval, respectively. By the formulas given in Refs. [11, 12], *R*(*i*) and *d*(*i*) can be be defined by

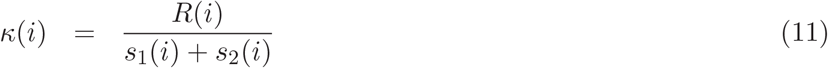

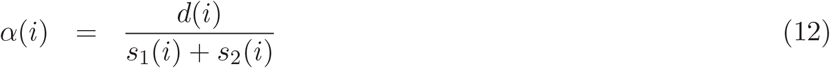

Since there is no information on recovered asymptomatic infected individuals, we take

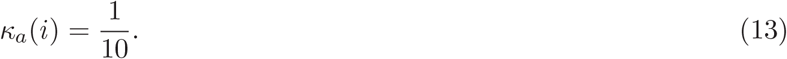

That is, an asymptomatic infected individual will recover in average 10 days. The calculated *κ*(*i*)′*s* and *α*(*i*)′*s* are shown in Table 1.

**Table 1.** The data of the first COVID-19 epidemic in Shanghai of SARDDE. Where NCSII represents the number [13] of current symptomatic infected individuals individuals. NRSII and NCDI represent the numbers [13] of calmative recovered symptomatic infected and died individuals, respectively.

| $i$ | Day | Date | NCSII | NRSII | NCDI | $\kappa(i)$ | $\alpha(i)$ | $\theta_1(i)$ | $\theta_2(i)$ |
| --- | --- | --- | --- | --- | --- | --- | --- | --- | --- |
| **1** | 0 | 1.20 | 2 | 0 | 0 | 0.018182 | 0 | 1 | 1 |
|  | 4 | 1.24 | 32 | 1 | 0 |  |  |  |  |
| **2** | 5 | 1.25 | 39 | 1 | 1 | 0.01108 | 0.0027701 | 0.334 | 0.2 |
|  | 10 | 1.30 | 122 | 5 | 1 |  |  |  |  |
| **3** | 11 | 1.31 | 143 | 9 | 1 | 0.0096805 | 0 | 0.173 | 0.189 |
|  | 16 | 2.05 | 238 | 15 | 1 |  |  |  |  |
| **4** | 17 | 2.06 | 243 | 25 | 1 | 0.024731 | 0 | 0.045 | 0.17 |
|  | 24 | 2.24 | 255 | 62 | 1 |  |  |  |  |
| **5** | 25 | 2.13 | 235 | 90 | 1 | 0.11083 | 0.0010076 | 0.009 | 0.173 |
|  | 43 | 3.03 | 41 | 294 | 3 |  |  |  |  |
| **6** | 44 | 3.04 | 37 | 298 | 3 | 0.13568 | 0 | 0 | 0 |
|  | 52 | 3.12 | 14 | 321 | 3 |  |  |  |  |
| **7** | 53 | 3.13 | 11 | 324 | 3 | 0.061538 | 0.015385 | 0 | 0 |
|  | 70 | 3.30 | 7 | 327 | 5 |  |  |  |  |
| **8** | 71 | 3.31 | 6 | 327 | 6 | 0.054945 | 0.021978 | 0 | 0 |
|  | 103 | 5.03 | 0 | 332 | 7 |  |  |  |  |

Second it needs to determine the parameters 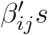 in the SARDDE. One can assume that *S* = 1 because the effects of *S* can be deleted by calculated 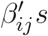. This makes the calculated 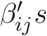 have general sense. Using the practical data of the first COVID-19 epidemic in Shanghai [13](also see the second line in Table 1) selects following initial condition:

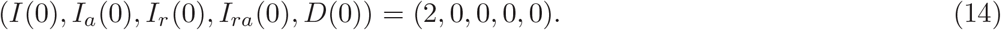

Substitute parameters *κ*(1), *α*(1), *θ*_1_(1) and *θ*_2_(1) listed in Table 1 into system (1). Using a minimization error square criterion:

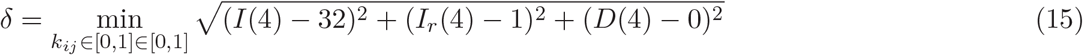

determines 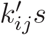.

A group (*β*_11_, *β*_12_, *β*_21_, *β*_22_) that makes *δ* be “smallest” (considering continued simulations) are

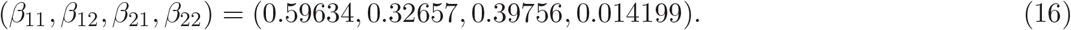

The first 4 days’ simulations of system (1) with the above equation parameters are shown in Figs. 3(a) and 3(b). The simulation results are in good agreement with the reported clinical data (see the first solid and dash lines and legends in Figs.2(a) and (b)).

Third it needs to determine: *θ*_1_(*i*), *θ*_2_(*i*), *i* = 2, 3, …, 8. Denote

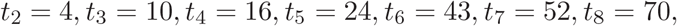

and *D*_*s*_(*t*_*i*_) and *D*_*sr*_(*t*_*i*_) to be the numbers of the Shanghai CONVID-19 current symptomatic infected and recovered individuals at *t*_*i*_, respectively, and *D*_*c*_(*t*_*i*_) the cumulative died individuals at *t*_*i*_.

Using the minimization error square criterion:

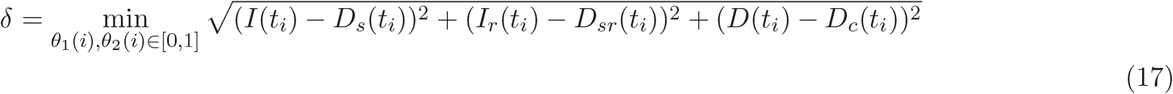

determines the *θ*_1_(*i*) and *θ*_2_(*i*). The calculated results are shown in Table 1.The corresponding simulation results of system (1) are shown in Fig.3(a) and 3(b). Observe that the simulation results of SARDDE model (1) describe well the dynamics of the first COVID-19 epidemic in Shanghai.

### 3.2 Discussions, virtual simulations

1. On the days 0, 4, 10, 16 24 and 103, the numbers of practical and simulated current symptomatic individuals are approximately the same. On the days 43, 52 and 70 they have 5, 3 and 2 differences, respectively.
2. On the days 0, 4, 10 and 16, the numbers of practical and simulated cumulative recovered symptomatic individuals are approximately the same, respectively. On the day 103, it has only 1 difference. On the days 24, 52 and 70, they have only 2 differences. On the day 43 it has 5 differences.
3. The all numbers of practical and simulated cumulative died individuals are approximately the same on the days 0, 4, 10, 16, 24, 43, 52, and 70. On the day 103, it has only 1 difference.
4. There is no information on current symptomatic infected and recovered symptomatic infected individuals. But it has reported that after the 70th day, March 30, there is no symptomatic infected individuals until the 110 day, May 9 [13]. Our simulation results shows that on the 70 day, the number of the simulated current symptomatic infected individuals were approximately to be 5, which can be explained that the 5 persons were not in hospitals and monitored.
5. Computed results (see (16)) show that the ratio of the transmission rate of asymptomatic and symptomatic individuals infecting susceptible population to become symptomatic individuals is about 0.54762 (*β*_21_: *β*_11_). This suggests that asymptomatic individuals cause lesser symptomatic spread than symptomatic individuals do.
6. The computed results (see (16)) also show that the ratios of the transmission rates of asymptomatic and symptomatic individuals infecting susceptible population to become asymptomatic and symptomatic individuals are about 0.67 (*β*_12_: *β*_11_) and 0.043 (*β*_22_: *β*_21_), respectively. This suggests that both symptomatic and asymptomatic individuals cause lesser asymptomatic spreads than symptomatic spreads.
7. The criterions (7) and (8) of the asymptotical stability of the disease-free equilibrium of SARDDE at eight time intervals are listed in the 5th ^ 8th columns in Table 2. It shows that the blocking rates (1 − *θ*_1_, 1 − *θ*_2_) reach to (95.5%, 83%) cannot prevent the transmission of the COVID-19 epidemic. The blocking rates (1 − *θ*_1_, 1 − *θ*_2_) reach to (99.1%, 82.7%) the disease-free equilibrium becomes asymptotical stability.

**Table 2.** The criterions of the asymptotical stability and disease spreading of the disease-free equilibrium of SARDDE at eight time intervals.

| $i$ | Day | $\theta_1(i)$ | $\theta_2(i)$ | $a_{11} + a_{22}$ | $a_{12}a_{21} - a_{11}a_{22}$ | $S_p^1$ | $S_p^2$ |
| --- | --- | --- | --- | --- | --- | --- | --- |
| 1 | 0 | 1 | 1 | 0.49236 | 0.17944 | 9.508 | 5.9917 |
| 2 | 5 | 0.334 | 0.2 | 0.088167 | 0.026679 | 2.3458 | 1.9873 |
| 3 | 11 | 0.173 | 0.189 | -0.0038301 | 0.013343 | 2.0153 | 1.5766 |
| 4 | 17 | 0.045 | 0.17 | -0.095482 | 0.0011986 | 0.30632 | 0.9476 |
| 5 | 25 | 0.009 | 0.173 | -0.20401 | -0.010183 | 0.052035 | 0.26673 |
| 6 | 44 | 0 | 0 | -0.23568 | -0.013568 | 0 | 0 |
| 7 | 53 | 0 | 0 | -0.17692 | -0.0076923 | 0 | 0 |
| 8 | 71 | 0 | 0 | -0.17692 | -0.0076923 | 0 | 0 |

Now assume that after the day 24, February 12, it still keeps the blocking rates (*θ*_1_(4), *θ*_2_(4)), the cure rates (*κ*(4), *κ*_*a*_(4)), and the died rate *α*(4) until the day 103, May 2. The simulation results of SARDDE are shown in Figs 4(a) and 4(b). Observe that the numbers of the current symptomatic and asymptomatic infected individuals reach to about 609 and 379, respectively. The numbers of cumulative recovered symptomatic and died individuals reach to about 855 and 1, because the died rate *α*(4) = 0.

**Figure 4:**
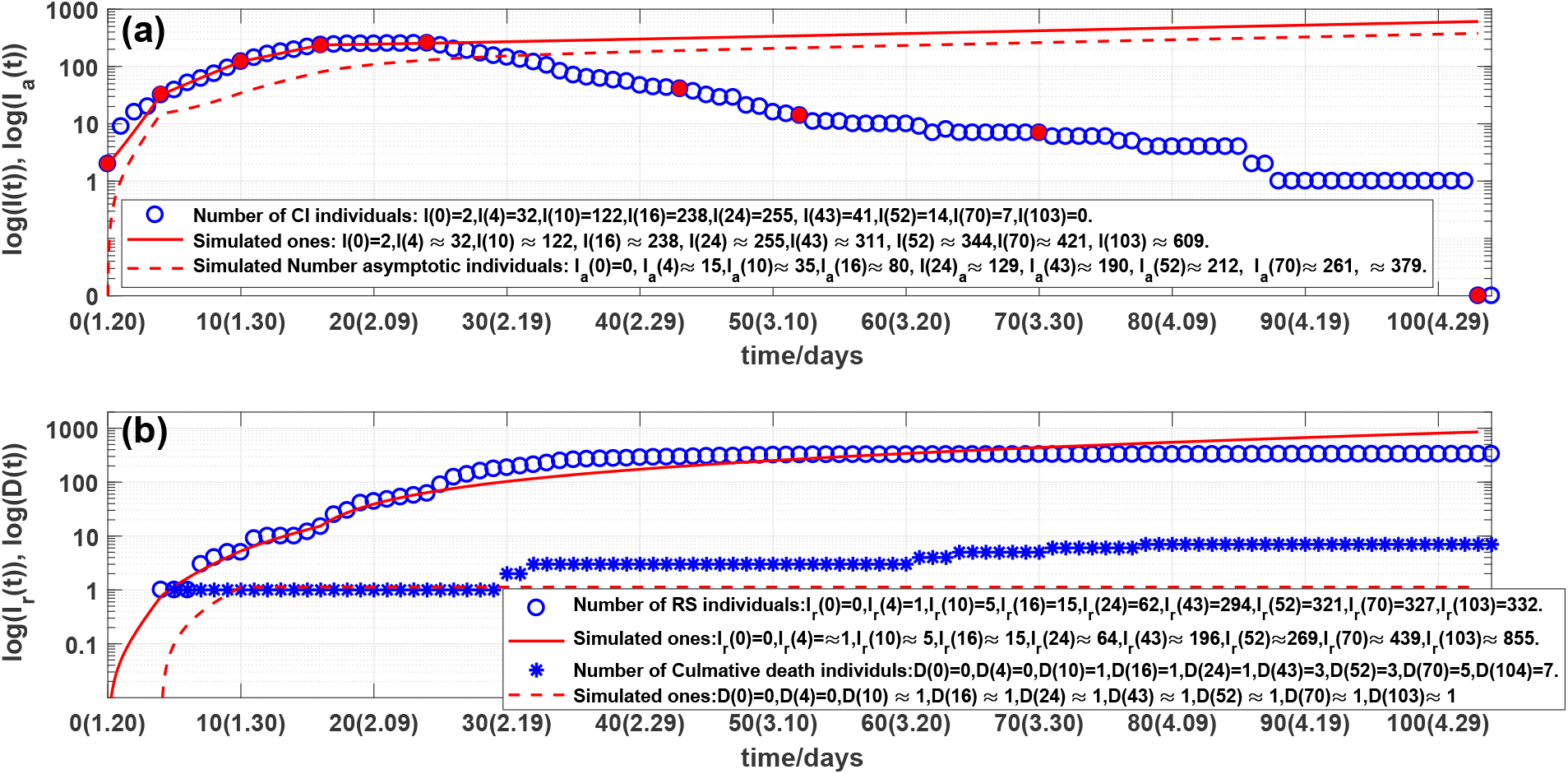
Outcomes of the numbers of:(a) current symptomatic individuals, representing by circles. Solid line and dash line are the stimulated current symptomatic and asymptomatic individuals of system (1); (b) cumulative recovered symptomatic and died individuals, representing by circles and stars, respectively. Solid line and dash line are corresponding simulations of system (1).

Furthermore assume that after the day 43, May 3, it still keeps the blocking rates (*θ*_1_(5), *θ*_2_(5)), the cure rates (*κ*(5), *κ*_*a*_(5)), and the died rate *α*(5) until the day 103, May 2. The simulation results of SARDDE are shown in Figs 5(a) and 5(b). Observe that the numbers of the current symptomatic and asymptomatic infected individuals are both less than one, respectively. The numbers of cumulative recovered symptomatic and died individuals are about 338 and 4, respectively. The results suggest that using the data before the day 43 (about 19 days after the turning point) can approximately estimate the following outcome of the first COVID-19 academic in Shanghai.

**Figure 5:**
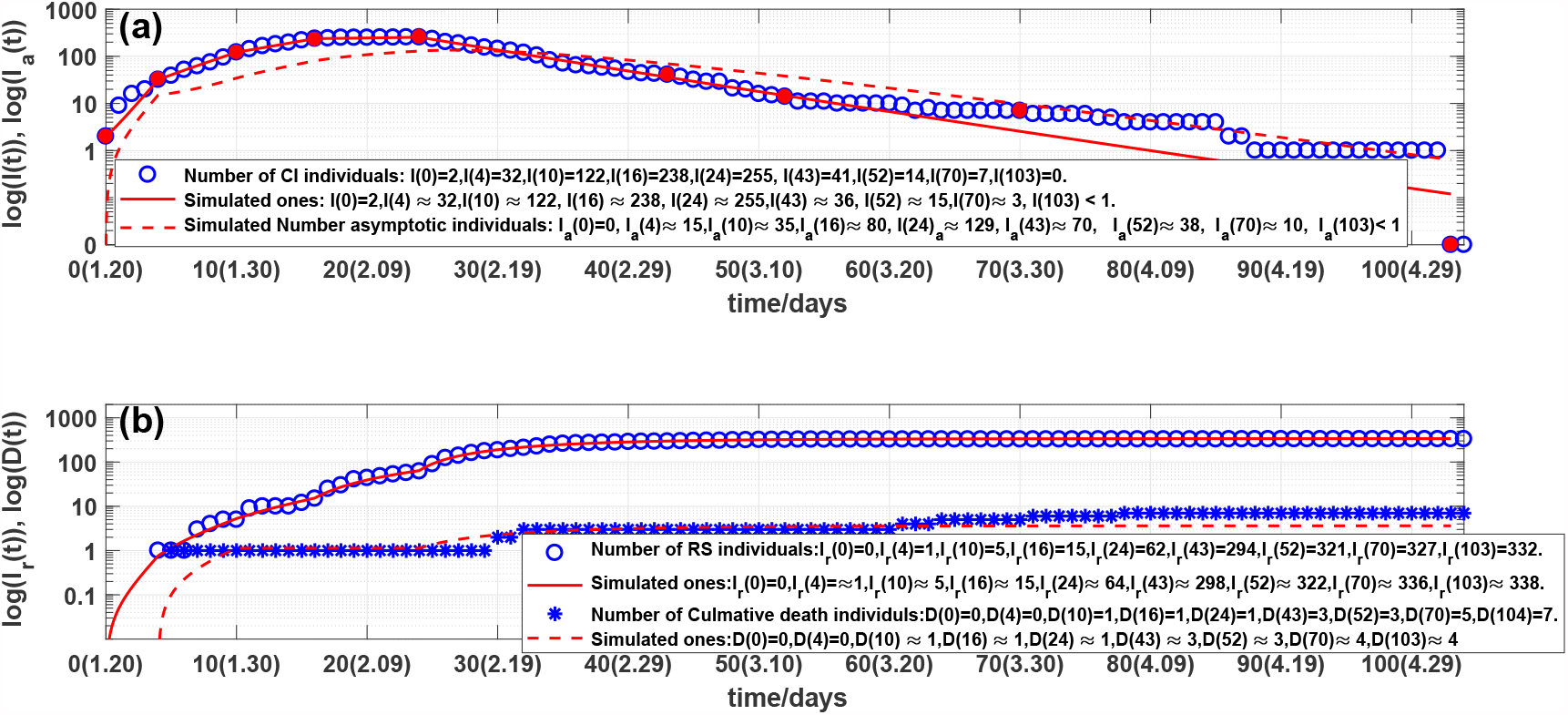
Outcomes of the numbers of:(a) current symptomatic individuals, representing by circles. Solid line and dash line are the stimulated current symptomatic and asymptomatic of system (1); (b) cumulative recovered symptomatic and died individuals, representing by circles and stars, respectively. Solid line and dash line are corresponding simulations of system (1).

In summary, SARDDE model (1) can simulate the outcomes of the first COVID-19 epidemic in Shanghai. The calculated equation parameters can help us to understand and explain the mechanism of epidemic diseases and control strategies for the event of the practical epidemic.

## 4 Conclusions

The main contributions of this paper are summarized as follows:

1. Using one our previous model [11, 12] simulates the dynamics of the first COVID19 epidemic in Shanghai reported by [13]. The simulation results are in good agreement with the reported clinic data [13], particularly the long term’s prediction.
2. Our model is simpler than a previous one [8] and obtains better simulation agreements than our previous model [8].
3. Eight of the nine model parameters are determined by the practical clinic data with numerical simulations. Only assumption is on parameter *κ*_*a*_(*i*) = 1*/*10 because no asymptotical infected individuals were reported.
4. The selections of the transmission rates *β*_*ij*_ are difficult because different combinations of *β*_*ij*_ can produce very closed simulation errors (see (15)). Agreements of followed simulations are used to judge the reasonableness of the selected 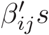.
5. The values (see (16)) of 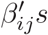 suggest that both symptomatic and asymptomatic individuals arise lesser asymptomatic infections. The symptomatic infections arising from the symptomatic individuals play an important role.
6. Complex prevent measures to symptomatic and asymptomatic infections can be represented by uniform forms *θ*_1_(*i*)′*s* and *θ*_2_(*i*)′*s*, respectively. Both theoretical analysis and numerical simulations show that the blocking rates to symptomatic and asymptomatic infected individuals reach to 95.5% (1 - *θ*_1_(4)) and 83% (1 - *θ*_2_(4)), the transmissions of the COVID-19 epidemic still continue.
7. Simulations showed that using the data form the beginning to the day after about 19 days the turning points, one can estimate well or approximately the following outcomes of the first COVID-19 academic in Shanghai.
8. The strict prevention and control strategies implemented by Shanghai government is not only very effective but also completely necessary.
9. There are a large numbers of articles on the modeling and prediction of COVID-19 epidemics. To our knowledge, no such a model was used to describe the dynamics of the first COVID-19 in Shanghai. It is expected that the presented researches can provide better understanding, explaining, and dominating for epidemic spreads, preventions and controls not only in Shanghai but also in other regions.

## Data Availability

All data cited in this article can be found in referece [13] (http://wjw.sh.gov.cn/)

